# Predicting gestational age at birth in the context of preterm birth from multi-modal fetal MRI

**DOI:** 10.1101/2024.02.17.24302791

**Authors:** Diego Fajardo-Rojas, Megan Hall, Daniel Cromb, Mary A. Rutherford, Lisa Story, Emma Robinson, Jana Hutter

## Abstract

Preterm birth is associated with significant mortality and a risk for lifelong morbidity. The complex multifactorial aetiology hampers accurate prediction and thus optimal care. A pipeline consisting of bespoke machine learning methods for data imputation, feature selection, and regression models to predict gestational age (GA) at birth was developed and evaluated from comprehensive multi-modal morphological and functional fetal MRI data from 176 control cases and 67 preterm birth cases. The GA at birth predictions were classified into term and preterm categories and their accuracy, sensitivity, and specificity were reported. An ablation study was performed to further validate the design of the pipeline. The pipeline achieves an R2 score of 0.51 and a mean absolute error of 2.22 weeks. It also achieves a 0.88 accuracy, 0.86 sensitivity, and 0.89 specificity, outperforming previous classification efforts in the literature. The predominant features selected by the pipeline include cervical length and various placental T2* values. The confluence of fast, motion-robust and multi-modal fetal MRI techniques and machine learning prediction allowed the prediction of the gestation at birth. This information is essential for any pregnancy. To the best of our knowledge, preterm birth had only been addressed as a classification problem in the literature. Therefore, this work provides a proof of concept. Future work will increase the cohort size to allow for finer stratification within the preterm birth cohort.

**Author summary:** Preterm birth is defined as the birth of a baby before the 37th week of pregnancy. It poses a serious risk to the life of a newborn and it is associated with a variety of severe lifelong health problems. Currently, the causes of preterm birth are not completely understood and therefore predicting when a baby will be born prematurely remains a challenging problem. Fetal MRI is an imaging technique that can provide detailed information about the development of the fetus and it is used to support the care of pregnancies at high-risk of preterm birth. Our work combines machine learning techniques with fetal MRI to predict gestational age at birth. The ability to predict this information is crucial for providing adequate care and effective delivery planning. The main contribution of our study is demonstrating that it is possible to make use of all the information obtained from fetal MRI to estimate the delivery date of a baby. To the best of our knowledge, this is the first study to combine machine learning with such a rich data set to produce these important predictions.

## Introduction

Preterm birth is defined as a live birth before 37 completed weeks of gestation [1]. It is estimated that every year 13.4 million babies are born prematurely, corresponding to a global preterm rate of around 9.9% [2]. Prematurity is the leading cause of mortality among children under 5 years accounting for 17.7% of the 5.3 million yearly deaths in this age group [3]. Complications associated with preterm birth are also the leading cause of neonatal mortality, accounting for 36% of these deaths [3]. The chances of survival of preterm babies are directly related to their gestational age (GA) at birth, with survival chances increasing from less than 18% for babies born at 22 weeks to over 95% for babies born at 29 weeks or later [4–6]. Despite advances in perinatal and neonatal care [4–9] survival critically depends on every additional week in-utero.

A continuous rise in survival rate has not translated into a decrease of the short- or long-term morbidity associated with preterm birth [8–10]. Short-term outcomes of premature birth include infections, bronchopulmonary dysplasia, retinopathy, necrotising enterocolitis, and brain disorders [11]. Long-term consequences include an increased risk of neuropsychiatric disorders such as psychosis, neurodevelopmental disabilities such as cerebral palsy and neuromotor dysfunction, adverse sensory outcomes such as hearing and visual impairment, as well as disabilities encompassing learning, cognition, and behaviour [10, 12, 13]. Similar to mortality rates, the incidence and severity of short- and long-term consequences of preterm birth are inversely related to GA at birth [11, 14, 15]. GA at birth is also correlated to social aspects later in life such as income and education level [15].

Reducing the incidence of preterm birth and the impact of its consequences would not only alleviate the burden on individual patients and their families, but also on entire healthcare systems, since the lifetime cost of preterm births in the USA (in 2016) was estimated to be $25.2 billion [16]. Unsurprisingly, a review of the literature on the economic consequences of preterm birth found a prevailing inverse relation between economic costs and GA at birth, regardless of methodology, date, or country of publication [17].

Preterm birth is classified into three subcategories: extremely preterm (less than 28 weeks), very preterm (28 to 32 weeks), and late preterm (32 to 37 weeks) [1], with further categorisation by clinical presentation: medically induced (or iatrogenic) and spontaneous [18]. While maternal and fetal indicators for iatrogenic preterm birth are well characterised and include conditions such as pre-eclampisa and fetal growth restriction (associated with 30.1% of cases) [19, 20], the aetiologies underlying spontaneous preterm birth are complex, varied, and poorly understood [21]. Causes include - but are not restricted to-infection or inflammation, vascular disease (leading to uterine ischaemia), uterine overdistention, and cervical injury. The latter can be a consequence of LLETZ procedures, cervical cone biopsies for abnormal smear tests, and injuries resulting from emergency C-sections in previous pregnancies [19, 22]. However, definitive causes are registered for only 50% [23, 24] of cases. As such, spontaneous preterm birth should more broadly be considered a syndrome resulting from multiple intricate causes [19, 25].

Despite this complexity, several risk factors have been identified [19, 21, 26] (see Table 1) and are useful, both to provide insights and to help identify at-risk women. The wide variety of factors thereby matches the aetiological complexity of preterm birth. Even within the same clinical subtype, some factors can have opposite effects. For example, low maternal body mass index (BMI) is a risk factor for fetal growth restriction but protective against preeclampsia, whereas these roles are reversed for maternal obesity [27].

**Table 1.** Most common risk factors for preterm birth [19, 21, 26].

| Risk Factors for Preterm Birth. |
| --- |
| African-American ethnicity |
| Depression |
| Family history of preterm birth |
| History of cervical excision |
| Infections (genitourinary or extragenital) |
| Low educational attainment |
| Low socio-economic status |
| Maternal age (low and high) |
| Maternal body mass index (low and high) |
| Multiple gestation (twins, triplets, etc) |
| Periodontal disease |
| Prior preterm birth |
| Stress |
| Stillbirth or induced abortion history |
| Tobacco use |
| Use of assisted reproductive technologies |
| Uterine anomalies |

Currently there are three leading indicators used in clinical practice to identify women at high risk. The strongest predictor is a history of previous preterm birth or cervical surgery or injury (32% chance of recurrent preterm birth) [19, 28]. The other two biomarkers are mid-trimester cervical length below 25mm [28, 29], measured via vaginal ultrasonography; and the presence of more than 50ng/mL fetal fibronectin, a glycoprotein usually absent in cervicovaginal fluid from 18 weeks of gestation and an indicator of choriodecidual disruption. The absence of any of these factors suggests the likelihood of delivering within the following 7 days is only around 1% [19, 28]. These factors have been combined within clinical practice to improve their predictive capabilities [30, 31]. In other analyses, the combination of these predictors also reduced the average cost of high-risk pregnancies. [32, 33].

Another modality with good potential to investigate preterm birth is Magnetic Resonance Imaging (MRI). Fetal MRI is used as a complementary modality to the commonly used ultrasound screening due to its higher resolution, operator independence and suitability for use on women with a higher BMI. It is also non-invasive with no evidence indicating any risk to the fetus or mother [34–36]. Another key advantage of MRI is that it offers multiple complementary contrasts that can support comprehensive functional evaluation of fetal and maternal tissues [37]. Available contrasts include T2-weighted anatomical imaging, T2* relaxometry (which provides an indirect measure of oxygenation [38]), diffusion MRI (which can quantify alterations in tissue microstructure [39–42]), flow measurements and T2 relaxometry. Past studies have largely focused on individual organs such as investigating changes to lung [43], thymus volumes [44], or assessing placental microstructure by measuring T2* and ADC values [45]. One study measured umbilical vein T2 values as a potential marker of intrauterine growth restriction [46]).

While predictive machine learning (ML) models have enjoyed an ever-increasing popularity, preterm birth has only been addressed as a classification problem. Models based on electronic health records, uterine electromyography and transvaginal ultrasound [47] reported accuracies of approximately 0.77 [48–50], while studies based on electrohysterography reported values above 0.94 [51–53], with the latter, however, only including records of women with recorded contractile activity [53, 54].

Machine learning applied to structural MR measurements has been successful at predicting GA at the time of the scan during pregnancy. For example, Convolutional Neural Networks trained on fetal brain MRI have been able to outperform current clinical methods to estimate GA at the time of scan [55, 56]. A different study managed to obtain a mean absolute error of 6.1 days by developing bespoke features from 3D ultrasound and using a regression forest for prediction [57].

For this work, a stacking approach was chosen to predict GA at the time of birth. Stacking is an ensembling technique that consists of combining the predictions of individual base models by training a meta-model [58]. Stacking was introduced by Wolpert in 1992 to improve the predictions and generalisability of individual classification models [59]. In 1996 Breiman [60] showed that stacking was also suitable for regression problems, while in 1999 Ting and Witten [61] generalised the technique further by stacking three different types of base models and exploring different meta-models than the ones used in previous work. Ensemble methods such as stacking have the statistical advantage of reducing the risk of overfitting to the training data by taking into account the predictions of all the base models, as well as the representational advantage of expanding the space of available models by combining the base models into meta-models [62].

In recent years, stacking has been successful at various tasks such as genomic prediction [63], protein interactions prediction [64], or prostate cancer detection [65]. These works take advantage of more recent ML learning models, e.g., Yi et. al. [64] use Support Vector Machines and XGBoost models as part of their base models, while Wang et. al. explore using a Random Forest as their meta-model [65].

The present study combines a uniquely rich MR data acquisition including both anatomical and functional scans of multiple fetal organs, and multimodal MRI of the placenta, with a ML pipeline based on stacking. To the best of our knowledge, this is the first work to leverage the advantages of stacking methods together with a comprehensive multi-modal data set to predict GA at birth.

## Methods

This section contains a detailed outline of the development of the ML pipeline introduced in this work. The pipeline was designed to address the challenges presented by the data. These include: a large number of derived features relative to the number of training examples, data imbalance, and missing data. These problems were addressed through feature selection, balanced training, and feature imputation. Throughout the development of the pipeline, different design options were investigated including changing data threshold for imputation, and models for feature selection and regression. The end product is a meta-model where predictions are stacked to obtain a final predicted GA at birth. The last subsection describes an ablation study, which investigates the impact of each component. Fig 1 illustrates the workflow of the project. The reader is invited to refer to it repeatedly to complement the description that follows.

**Fig 1.**
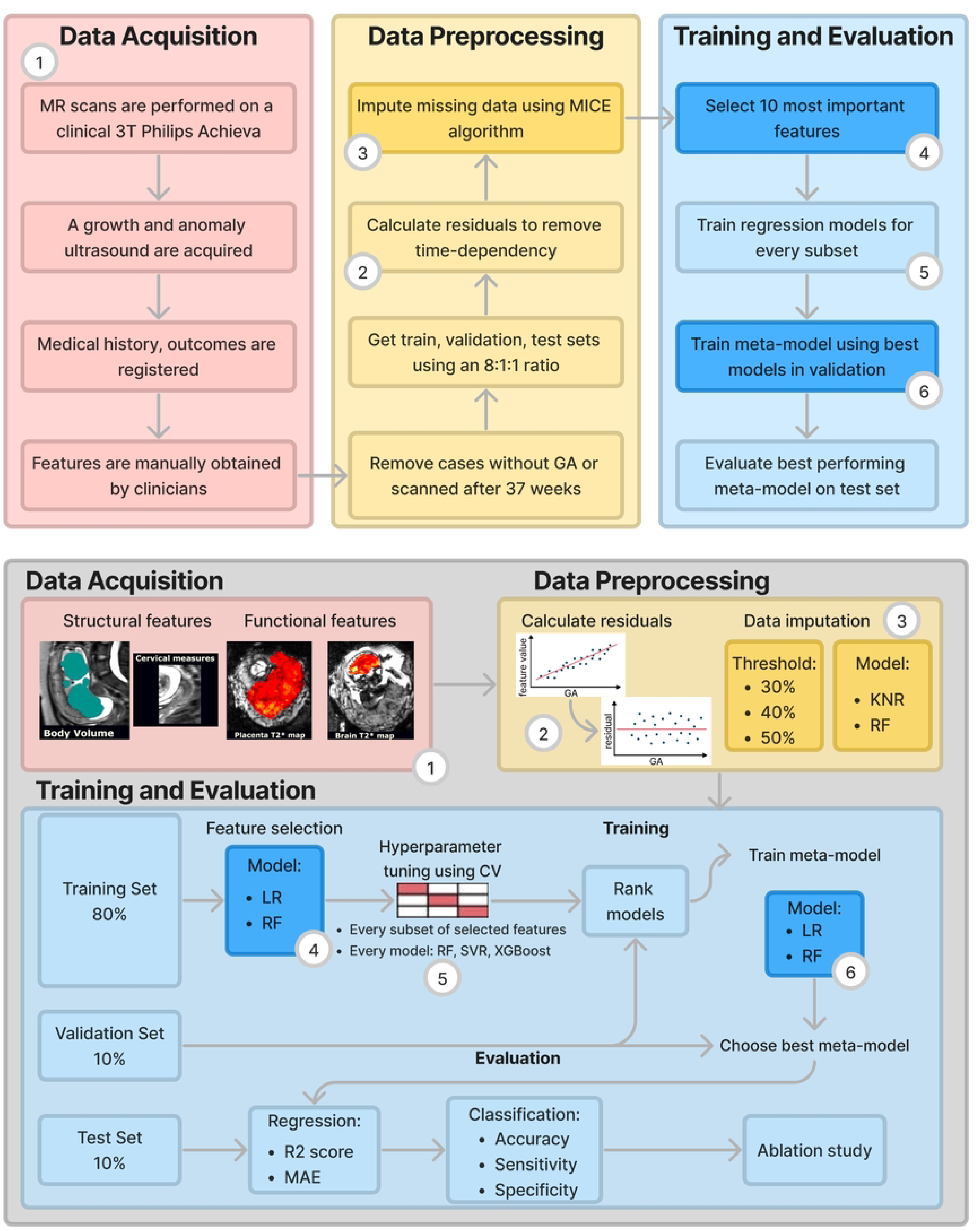
Schematic representation of the project pipeline. The boxes in a darker shade denote steps of the pipeline where different design options were explored. The top part of the figure shows the flow of the pipeline with fixed designed options, while the bottom part explicitly indicates the different design choices available for each step.

### Data

The data set used for this work comprises clinical records, MR data, and parameters manually extracted from ultrasound from 313 singleton pregnancies, acquired as part of four ethically approved studies: 14/LO/1169 (Placenta Imaging Project, Fulham Research Ethics Committee, approval received September 23, 2016), 19-SS-0032 (Inflammation study in pregnancy, South East Scotland Ethics Committee, approval received March 7, 2019), 21/WA/0075 (Congenital Heart Imaging Programme, Wales Research Ethics Committee, approval received March 8, 2021), and 21/SS/0082 (Individualised Risk prediction of adverse neonatal outcome in pregnancies that deliver preterm using advanced MRI techniques and machine learning, South East Scotland Ethics Committee, approval received March 2022). Informed consent was obtained in all instances.

From the 313 cases originally considered, 59 cases were excluded as they were lacking GA at delivery. We also removed 11 cases scanned after 37 weeks, since - in the context of predicting preterm birth - these would bias training of the model. This resulted in a final data set of 243 cases (see S1 Fig).

Recruitment for all the considered studies was opportunistic, with two studies particularly recruiting women at high risk of preterm birth based on obstetric history, ultrasound and biomarker findings. However, the stated difficulty in accurately predicting preterm birth renders this task difficult, and as a result recruitment and thus the data set available is biased towards term birth.

In all experiments data was split into train, validation, and test sets with an 8:1:1 ratio, keeping an equal proportion of term and preterm birth cases in each set.

### Image Acquisition and Processing

Imaging protocols were similar for each study: MR scans were performed on a clinical 3T Philips Achieva scanner between 15 and 40 weeks of gestation using a 32-channel cardiac coil (as is standard process for fetal imaging). For maternal comfort, padding was provided, imaging time was limited to under 90 minutes, and there was frequent verbal interaction and monitoring of heart rate and blood pressure. The protocol included both anatomical T2-weighted imaging and functional MR sequences. For the work presented here only T2* relaxometry among the functional sequences was used.

Anatomical information was acquired with a 2D multi-slice Turbo-Spin-Echo sequence in four to ten planes covering the fetal brain and uterus. Next, to allow for image-based shimming on the 3T scanner, a map of the B0 field was obtained; then shimming was performed for the organ of interest. Afterwards, functional MRI of the entire uterus was performed in coronal orientation using free-breathing multi-echo Gradient Echo with Echo Planar read-out.

In addition to the MRI, two ultrasound scans were performed: an anomaly scan (clinically performed between 19 and 21 weeks of gestation) and a second growth scan (including Doppler ultrasound) that was generally performed within one week of the MRI. In both cases, morphological measurements were manually extracted including abdominal and head circumference, bi-parietal diameter (i.e. the cross-sectional diameter of the skull), femur length, and expected weight. From the growth scan blood flow pulsatility indices were also estimated for the umbilical, uterine and mid-cerebral arteries.

The obtained MR images were processed to obtain quantitative values. For the anatomical data, slice-to-volume reconstruction [66] and learning-based segmentation [67] were applied separately for the brain, body and the placenta. Then regional volumes were calculated.

From the functional data, no motion correction was applied, since all echos for each slice were acquired within 200ms. Using the method described in [45], quantitative T2* values were obtained by fitting the signal of data from subsequent echo times for the entire uterine field of view (FOV). Values over 300ms were clipped to limit partial volume effects following common practise. Segmentation of the placenta, brain and lungs was performed manually. From these segmentations, regional volumes were calculated, as well as the mean, kurtosis, and skewness of their T2* distributions. These data acquisition steps are represented by index 1 in Fig 1.

## Summary of Derived Features

In addition to imaging derived features, demographics, obstetric and medical history of the patients (including previous pregnancies, miscarriages and preterm births) were recorded as well as any relevant information from the current pregnancy such as diagnosis of pre-eclampsia, gestational diabetes, fetal growth restriction or any other fetal or maternal pathology. Finally, the outcomes of the pregnancy were obtained, including gestational age at birth, birth weight-centile and any occurrence of major complications. Collectively the full set of features used by our models was summarised as follows (see S1 Table for more details):

1. **Clinical variables**: e.g. number of previous preterm deliveries, and maternal body mass index.
2. **Structural MRI metrics**: describing sizes of structures e.g. volumes of different brain regions or bi-parietal diameter of the foetal head; these were extracted from both the anatomical and functional scans
3. **Functional MRI metrics**: statistics derived from T2* distributions of the placenta, brain and lungs.
4. **Ultrasound metrics**: from both anomaly and growth ultrasounds - manually extracted by a trained sonographer e.g. the fetal head circumference, femur length.

### Feature cleaning

Prior to training it was vital to address the confound effect of gestational age at scan, as well as address the impact of missing data.

### Deconfounding

While GA at scan is a feature that would normally be available in a clinical setting, its impact on any learning model could lead to data leakage (e.g. by acting as a lower bound for GA at delivery). Moreover, as all features change dramatically with age [43, 44, 68, 69], it is necessary to disentangle the dominant effect of GA from more subtle signatures that might robustly predict preterm birth. For these reasons, GA was linearly regressed from all features using the method of internally studentised residuals [70]. See index 2 in Fig 1.

### Data Imputation

There was significant heterogeneity in the availability of features across the data set. Fetal and maternal motion, maternal discomfort, and clerical errors led to loss of data, with different features available for each of the 243 cases. For this reason, a regression-based approach to imputation, known as Multivariate imputation by chained equations (MICE) [71], was investigated (see S1 Algorithm). Following guidance from the literature, ten iterations of the model were performed [71, 72], with two different regression models: weighted *K*-Nearest Neighbours (KNR) [73] and Random Forests [74]. Both models were implemented in the standard way using Sci-kit Learn [75]. Imputation should not be applied to features with arbitrarily large amounts of missing data [72, 76]. Thus the impact of discarding features with more than 30%, 40%, or 50% missing values was investigated (see S2 Table for the missing percentages of each feature). Features with a greater percentage of missing values than the respective threshold were discarded. All remaining features were normalised (mean 0, std 1) afterwards.

Data imputation corresponds to index 3 in Fig 1. The boxes corresponding to this step are emphasised by a darker shade to represent that different options were investigated as part of the pipeline design process. The top part of the figure shows the flow of the pipeline with fixed design choices (e.g. if the choice is made to investigate a pipeline using a RF within the MICE algorithm to impute features with less than 40% of missing data). Conversely, the bottom part of the figure explicitly indicates the design choices that were investigated for this step.

### Training

Training was performed using a stacking approach in which a number of different classes of machine learning model were trained and these were ensembled together through the training of a meta model [59]. Base models consisted of: Random Forests (RF) [74], Support Vector Regression (SVR) [77], and XGBoost [78]. Each was chosen due to unique strengths: RF are interpretable and robust to overfitting [79]; SVR are robust to outliers and well-suited to small data sets [80, 81]; XGBoost offers state-of-the-art performance from sparse data sets [78]. Importantly they are all capable of capturing non-linear relationships but approach regularisation in different ways [80]. This suggests that they will perform differently on boundary cases, to produce diverse predictions that could benefit from ensembling.

Since a key challenge of training models on our data set has been the high number of features relative to examples (see S1 Table), feature selection was also performed to discourage overfitting. Two simple models were explored: linear regression and Random Forests. For each model trained, 10 features were selected. These two different design options are indicated by the boxes with a darker shade with index 4 in Fig 1.

Models were trained using the Sci-kit learn framework [75], with hyperparameters (see S3 Table) optimised using 3-fold cross-validated grid search [82]. The metric used for optimisation was the coefficient of determination (*R*^2^) [83]. Given fixed design choices on the previous steps, training was carried out every non-empty subset of the selected features. Since there are 1023 non-empty subsets of the ten selected features and 3 regression models, 3069 different regression models were trained in total (Index 5 in Fig 1). These were then composed via the training of a meta-model, for which two different methods were explored: Linear Regression and Random Forests (index 6 in Fig 1). Meta-models were trained on the *m* best performing base models, as validated through their *R*^2^ score on the validation set. The value of *m* was also optimised using the validation set.

### Ablation Study

An ablation study was conducted to validate the design of the proposed pipeline, with results compared against the best performing meta-model. Since XGBoost models may be trained with incomplete data, and without variance normalisation of the features (since the base learners are decision trees) the first two experiments consist of a single XGBoost model trained on unnormalised data. All experiments are described as follows:

1. Out-of-the-box XGBoost: XGBoost without preprocessing.
2. XGBoost with deconfounding: one XGboost was trained after linear deconfounding of features.
3. Imputation: all base predictive models were trained with deconfounding and imputation (using the imputation approach used in the best meta-model), without performing any upsampling or feature selection.
4. Correcting data imbalance: base models were trained with imputation and upsampling preterm cases in the training set.
5. Combining feature selecting with upsampling: This equates to evaluating the best performing base model, obtained without ensembling.
6. Meta-model without upsampling: the impact of upsampling in the whole pipeline was explored by turning it off. This is equivelent to the final meta-model without upsampling.
7. Meta-model: Reporting the performance of the proposed meta-model - obtained from the whole pipeline.

## Results

### Data Exploration

Table 2 shows key demographics, clinical information, and outcomes, divided into preterm and control cohorts. Specifically, the data set consisted of 176 control cases and 67 preterm cases. The distribution of the data according to the four temporal categories was 72.4% term, 14.8% late preterm, 7% very preterm, and 5.8% extremely preterm. Fig 2 shows the distribution of the five continuous features and outcomes included in Table 2, namely GA at scan, maternal BMI at scan, maternal age, GA at birth, and birth weight centile. The pairwise relationship between these is also plotted. For a statistical summary of the data set see S2 Table.

**Fig 2.**
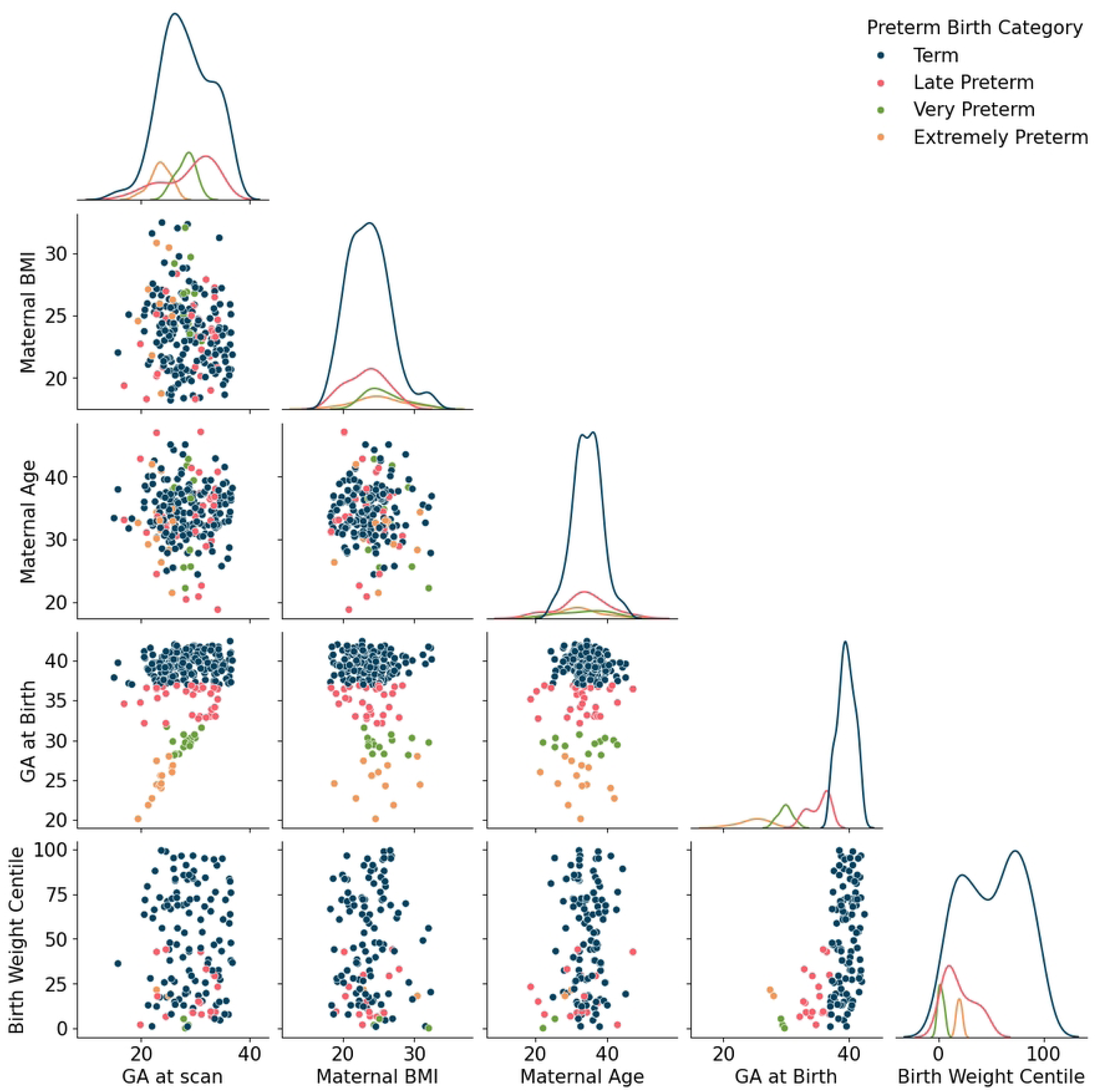
Data exploration. Distribution and pairwise relationship between GA at scan, maternal BMI at scan, maternal age, GA at birth, and birth weight centile in the data set.

**Table 2.** Key demographics, clinical information, and outcomes of participants.

|  | Preterm birth cohort | Control cohort |
| --- | --- | --- |
| Current pregnancy |  |  |
| Gestational age at scan [weeks] | 27.62±4.29 [16.86,34.14] | 28.39±4.65 [15.00,36.86] |
| Maternal BMI at scan [kg/m2] | 24.26±3.06 [18.31,32.05] | 23.59±2.94 [18.21,32.46] |
| Maternal age at scan [years] | 33.30±6.37 [18.82,47.17] | 34.50±3.92 [24.43,45.13] |
| Obstetric history |  |  |
| Previous preterm birth | 16.42% | 8.52% |
| Outcome |  |  |
| Gestational age at birth | 31.67±4.48 [20.14,36.86] | 37±42.43 [39.53,1.32] |
| Birth weight centile | 31.07±32.32 [0.00,95.53] | 53.23±28.52 [0.82,99.77] |
| Fetal sex | 54.69% female, 45.31% male | 55.56% female, 44.44% male |

Reported values are the mean ± SD in the case of continuous variables and percentages for discrete variables. The numbers in brackets are ranges.

### Meta-model

The best performing meta-model achieved an *R*^2^ score of 0.45 and a mean absolute eror (MAE) of 2.55 weeks on the validation set. This performance was achieved using the following settings in the pipeline. First, features with more than 50% missing values were discarded. Then, features with 50% or less missing values were imputed using the MICE algorithm with a KNR as its regression model and a RF was used for feature selection. In order of importance, the selected features were:

1. Cervical length measured from the sagittal plane of MR scan.
2. Mean whole placental T2* value measured from the MR scan.
3. End-diastolic flow measured from the growth ultrasound.
4. T2* brain to placenta ratio measured from the MR scan.
5. Bi-parietal diameter measured from the anomaly ultrasound.
6. Placental T2* kurtosis value measured from the MR scan.
7. Fetal head circumference measured from the anomaly ultrasound.
8. Brain T2* kurtosis value measured from the MR scan.
9. Estimated fetal weight at growth ultrasound.
10. Whole brain T2* volume value measured from the MR scan.

Fig 3 a) shows the mean decrease in impurity corresponding to each of these features. This is the metric used by Random Forests to assign importances to each feature [84].

**Fig 3.**
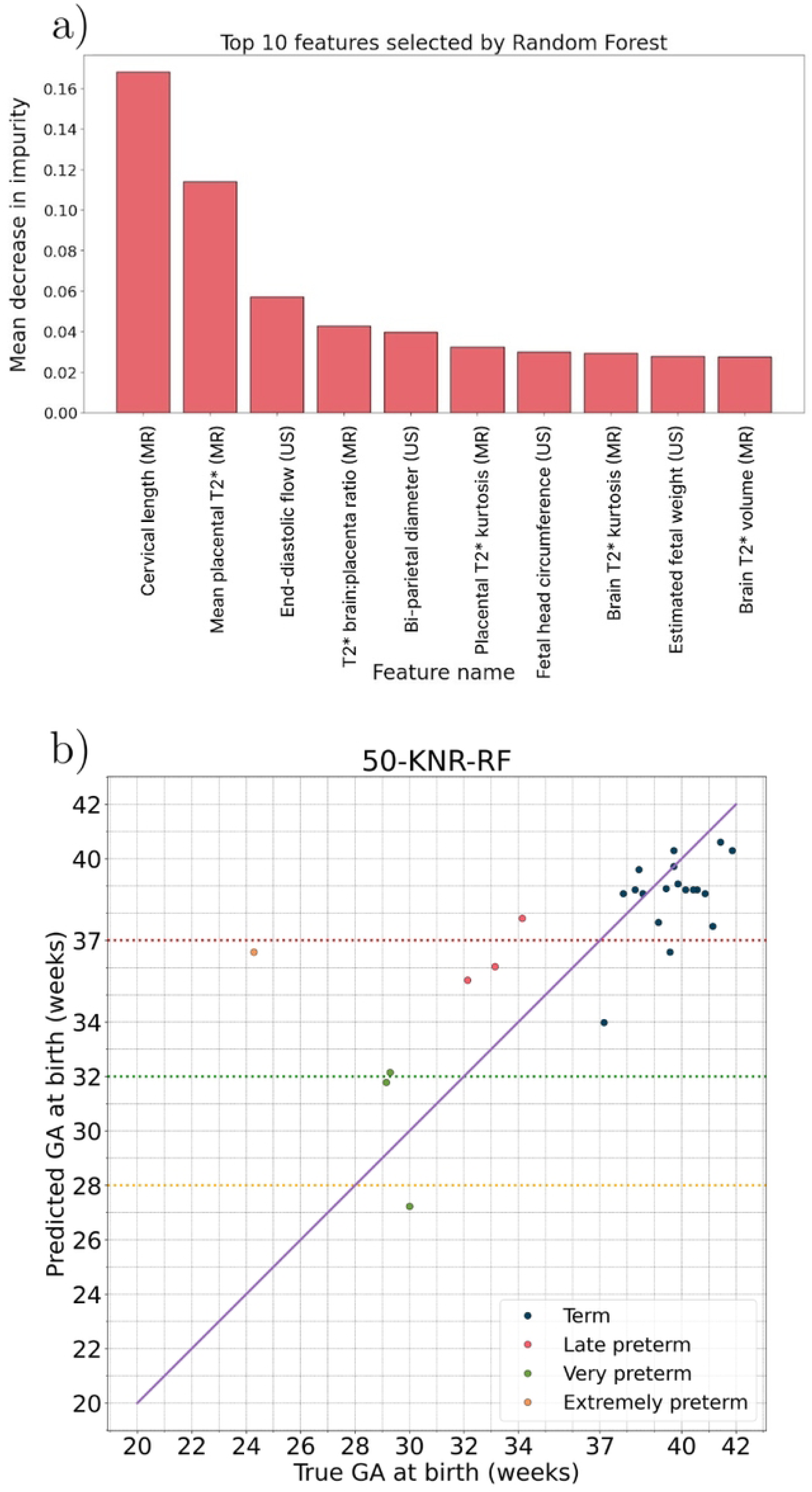
Feature importances and predictions of the meta-model. a) Feature importance score of the 10 most important features used to train the base models in the pipeline. b) Predictions made by the meta-model 50-KNR-RF on the test set, colourised according to their true preterm temporal category.

After training RF, SVR, and XGBoost models with every non-empty subset of these 10 features, the 14 models with the highest *R*^2^ score on the validation set were used as input for a RF meta-model. Specifically, the 14 base models that provided the features used by the meta-model were all SVRs, trained with different subsets of the selected features. In what follows, this meta-model will be referred to by abbreviating its components, i.e. 50-KNR-RF.

The metrics used for evaluation on the test set were the *R*^2^ score and the mean absolute error (MAE) measured in weeks. The cases were labeled as term (≥ 37 weeks) or preterm (< 37 weeks), according to the GA predicted by the meta-model, and accuracy, sensitivity, and specificity were also reported. On the test set 50-KNR-RF achieved an *R*^2^ score of 0.51 and a MAE of 2.22 weeks. After labeling each subject as term or preterm according to their predicted GA at birth, the meta-model achieved a 0.88 accuracy, 0.86 sensitivity, and 0.89 specificity. The predictions made by 50-KNR-RF on the test set are depicted in Fig. 3 b). It is worth noting that the model only misclassified one of the preterm cases and two term ones.

### Ablation study

The performance of each of the models resulting from the experiments of the ablation study is reported in Table 3. 50-KNR-RF outperformed every model in the ablation study in every evaluation metric. The best performing model within the experiments that consist of training different versions of the base models (experiments 3), 4), and 5)) was a SVR, which coincides with the type of models used as features for 50-KNR-RF.

**Table 3.** Evaluation of the models in the ablation study.

| Model | $R^2$ score | MAE | Accuracy | Sensitivity | Specificity |
| --- | --- | --- | --- | --- | --- |
| 1) Out-of-the-box XGBoost | -0.45 | 3.84 | 0.60 | 0.57 | 0.61 |
| 2) XGBoost with deconfounding | -0.34 | 3.61 | 0.64 | 0.43 | 0.72 |
| 3) Imputation, RF | 0.01 | 3.39 | 0.68 | 0.43 | 0.78 |
| 3) Imputation, SVR | 0.29 | 2.66 | 0.80 | 0.86 | 0.78 |
| 3) Imputation, XGBoost | 0.02 | 3.20 | 0.68 | 0.57 | 0.72 |
| 4) Correcting data imbalance, RF | 0.10 | 3.16 | 0.60 | 0.57 | 0.61 |
| 4) Correcting data imbalance, SVR | 0.34 | 2.75 | 0.76 | 0.86 | 0.72 |
| 4) Correcting data imbalance, XGBoost | -0.28 | 3.67 | 0.52 | 0.43 | 0.56 |
| 5) Combining feature selecting with upsampling, SVR | 0.15 | 3.48 | 0.72 | 0.86 | 0.67 |
| 6) Meta-model without upsampling, RF | 0.03 | 2.68 | 0.72 | 0.43 | 0.83 |
| 7) Meta-model, 50-KNR-RF | 0.51 | 2.22 | 0.88 | 0.86 | 0.89 |

## Discussion

Comprehensive multi-modal fetal data and ML models combine synergistically to predict GA at delivery. The developed pipeline acknowledges and addresses key challenges such as imbalances and missing features in the data set, both of which are common when investigating preterm birth.

As mentioned in the Introduction, preterm birth had so far only been addressed as a classification problem. The pipeline constructed in this work attempts to make more precise predictions by developing a regression model to predict GA at birth. In turn, these predictions can be classified as term or preterm to compare the performance of 50-KNR-RF to other classification models in the literature. Table 4 shows a comparison between 50-KNR-RF and the best performing models obtained by other recent studies. Out of the 5 models, 50-KNR-RF has the highest accuracy and specificity. The only model that achieves a higher sensitivity than 50-KNR-RF is one of the models developed by by Esty et al. [48]. However, the 0.93 sensitivity achieved by that model comes with the trade-off of having an accuracy and specificity lower than 0.73. In contrast, 50-KNR-RF achieved a score greater than 0.85 in every metric.

**Table 4.**
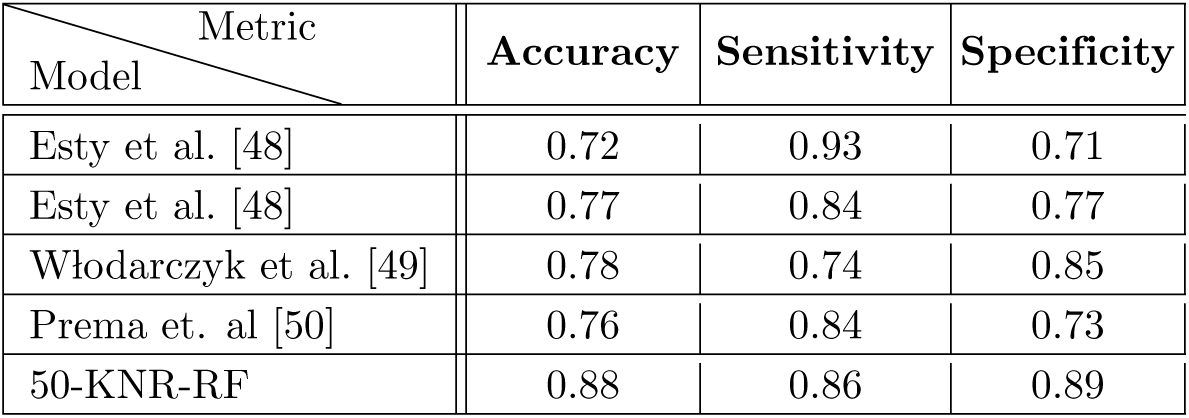
Classification performance of the meta-model obtained by this work and other models from recent studies.

To the best of our knowledge, the only other study on predicting GA at delivery using ML is the one by Heinsalu et al. [85], where they investigated models using a simpler version of the pipeline displayed in this work. Their best performing model achieves an *R*^2^ score of 0.66 and a MAE of 1.60, but their implementation suffers from data leakage at the imputation stage which makes these results unreliable. Nevertheless, the framework they established is valuable and served as the basis of the present work.

This study provides a proof of concept, but the clinical implementation of a reliable model that could predict the timing of delivery would have important benefits. These include ensuring women are transferred to appropriate neonatal care facilities. A timely transfer helps reduce neonatal mortality and decrease costs [37]. Another crucial example is the targeting of therapies to mitigate the effects of prematurity. Especifically, cortiscosteroids administration can help reduce intra-ventricular haemorraghe and promote lung maturity. The timing of this therapy is highly important, since it works best when administered within a week before delivery, and repeated doses increase the risk of adverse effects such as reduction in birthweight [86].

The results of the ablation study in Table 3 are helpful to understand the importance of every element of the pipeline. In experiments 1) and 2) it can be seen that even though XGBoost has a built-in mechanism to be trained using data sets with missing values, the results achieved with minimal preprocessing steps are among the worst in the study. In experiments 3) and 4) RF, SVR, and XGBoost models were trained using the data set obtained after imputation, without a feature selection selection step, and using upsampling in the case of experiment 4). On average, results in these experiments were better than in experiments 1) and 2) but upsampling did not translate in a significant difference between experiments 3) and 4). Contrastingly, the impact of upsampling in the whole pipeline can be appreciated by comparing the meta-model obtained in experiment 5) and 50-KNR-RF. The only difference between these two experiments is the use of upsampling during training. The sensitivity score of 0.43 obtained by the meta-model in experiment 5) evidences its inadequacy to address data imbalance, while the 0.86 score achieved by 50-KNR-RF shows that upsampling is effective in tackling this problem. Lastly, it can also be observed that 50-KNR-RF generalises well, i.e. it is not overfitting. 50-KNR-RF achieves an *R*^2^ score of 0.4530 and a MAE of 2.5489 weeks on the validation set and an *R*^2^ score of 0.5143 and a MAE of 2.2202 weeks on the test set. This is line with the literature, that suggests that ensemble models tend to generalise well [59]. The reason why the SVR model in experiment 6) has lower scores than its counterparts in experiments 3) and 4) in spite of being the best model prior to the ensembling of 50-KNR-RF is because it is not generalising well. It is the best performing model on the validation set, where it achieves an *R*^2^ score of 0.4878 and a MAE of 2.6418 weeks, but its performance on the test set is significantly worse. Combining the predictions of the 14 best models via 50-KNR-RF helps to solve this problem.

The features selected as the most important are in line with the literature. The importance of cervical length as a predictor in clinical practice is reflected by its consistent use in the models. Placental features obtained from MRI scans were other prominent features, which is in line with the current understanding of the mechanisms leading to iatrogenic preterm birth [45, 87]. The most common clinical indicator, number of previous preterm births, was not a predominant feature. This could be explained by the decision of approaching the problem as a regression instead of a classification one.

While this data set could be considered large given the comprehensive data acquisition, including a fetal MR scan in a cohort of women requiring a high level of medical care, its size is an important limitation for ML methods. The few examples of extremely preterm subjects available during training help explain the poor performance on the test set for this category. Taking into account the performance of 50-KNR-RF in the validation and test sets, the meta-model seems to generalise well. However, the size of the data set makes it hard to predict if the performance of the meta-model would generalise to new data.

Another limitation is the lack of information on the clinical presentation of preterm birth for every patient in the data set. Iatrogenic and spontaneous preterm births have different aetiologies and training separate models for each case could not only yield better predictions, but also help improve the understanding of each clinical presentation by differentiating their most predictive features. Future work will focus on such subgroups and on extracting relevant phenotypes associated with the different types of preterm birth.

Data obtained on a 1.5T and a 0.55T scanner were available for this study. However, these were not included as there is not a straightforward way to extrapolate the signals acquired by scanners with different magnetic field strengths [88]. Future experiments that include these types of data could test the adequacy of the elements of the pipeline, such as the method of internally studentised residuals, to make accurate predictions regardless of field strength.

There are other directions future research can take to expand or improve the methodology presented in this work. The implementation of the models is ready to benefit from larger or more complete data sets. Adding features known for their predictive power, such as quantitative fibronectin measurements, could improve the results. The performance of the meta-model demonstrates that structural and functional information obtained from MRI can be used to predict GA at delivery. An interesting direction is to make predictions directly from the images making use of deep learning techniques, bypassing the problem of missing data and the need of time-consuming measurements made by experienced clinicians. These techniques have been explored to classify preterm and term patients by automatic measurements of cervical length from transvaginal ultrasound [49] and to estimate GA at scan from fetal brain MRI [55, 56].

MRI remains an expensive modality, however, with an increasing use of fetal MRI, the pipeline presented in this study helps to address a question essential for any pregnancy, and can find an application regardless of the indication of the scan. One of the fundamental contributions of this work is that it shows that fetal MR data acquired as part of diagnostic care or research can be used to obtain useful predictions on the GA at delivery, which in turn can inform the care provided to all pregnancies.

## Data Availability

All data code and data are available online at https://github.com/dfajardorojas/ml-for-preterm-birth-

## Acknowledgments

The authors acknowledge the invaluable help of the radiographers and midwives while acquiring the data presented here.

## Author Contributions

**Conceptualization:** Diego Fajardo-Rojas, Emma Robinson, Jana Hutter.

**Data Curation:** Megan Hall, Daniel Cromb, Lisa Story.

**Formal Analysis:** Diego Fajardo-Rojas.

**Investigation:** Diego Fajardo-Rojas.

**Methodology:** Diego Fajardo-Rojas, Emma Robinson, Jana Hutter.

**Software:** Diego Fajardo-Rojas.

**Supervision:** Mary A. Rutherford, Lisa Story, Emma Robinson, Jana Hutter.

**Validation:** Diego Fajardo-Rojas.

**Visualization:** Diego Fajardo-Rojas.

**Writing – Original Draft Preparation:** Diego Fajardo-Rojas.

**Writing – Review & Editing:** Emma Robinson, Jana Hutter.

## Supporting information

**S1 Table.**
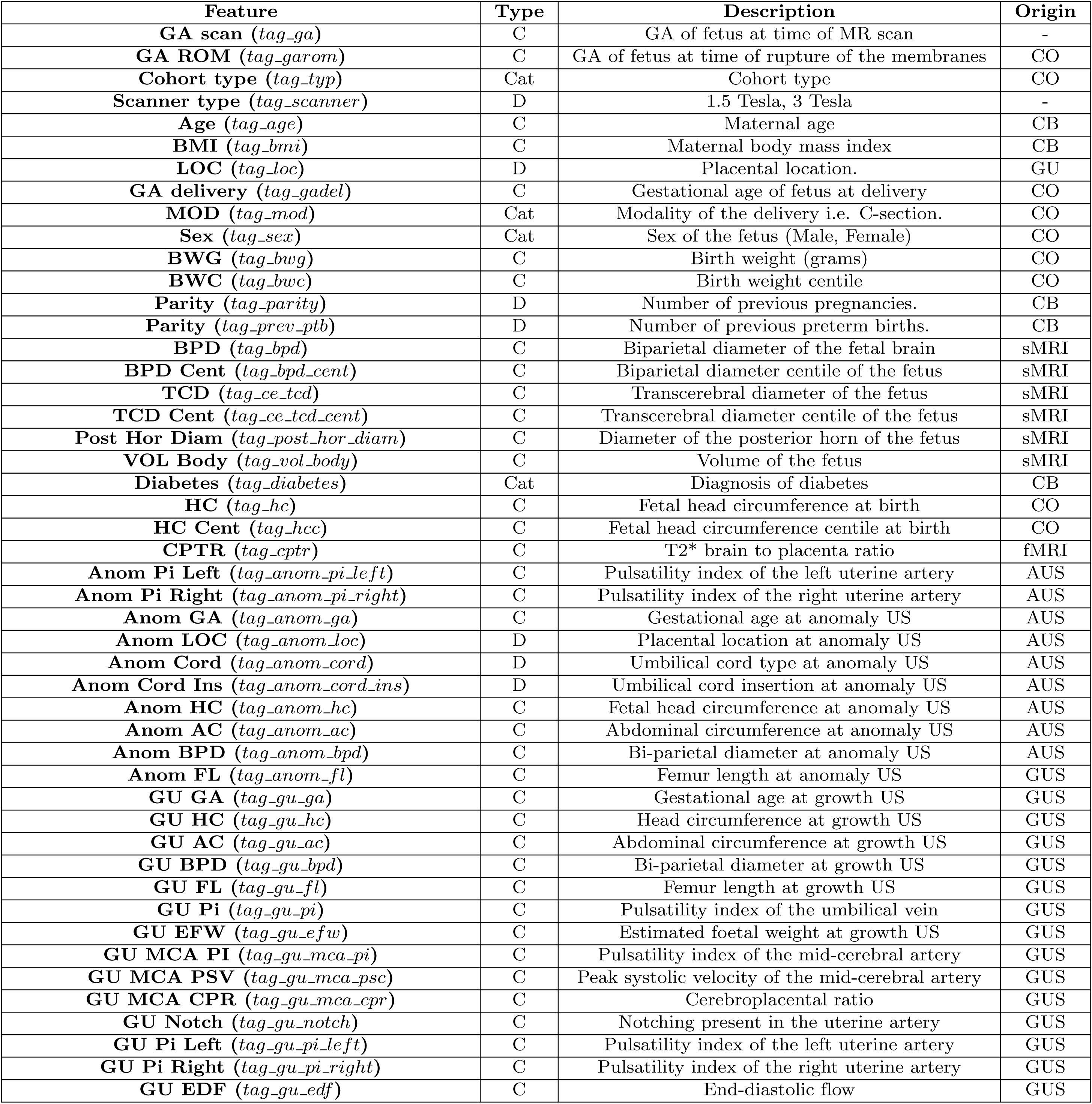
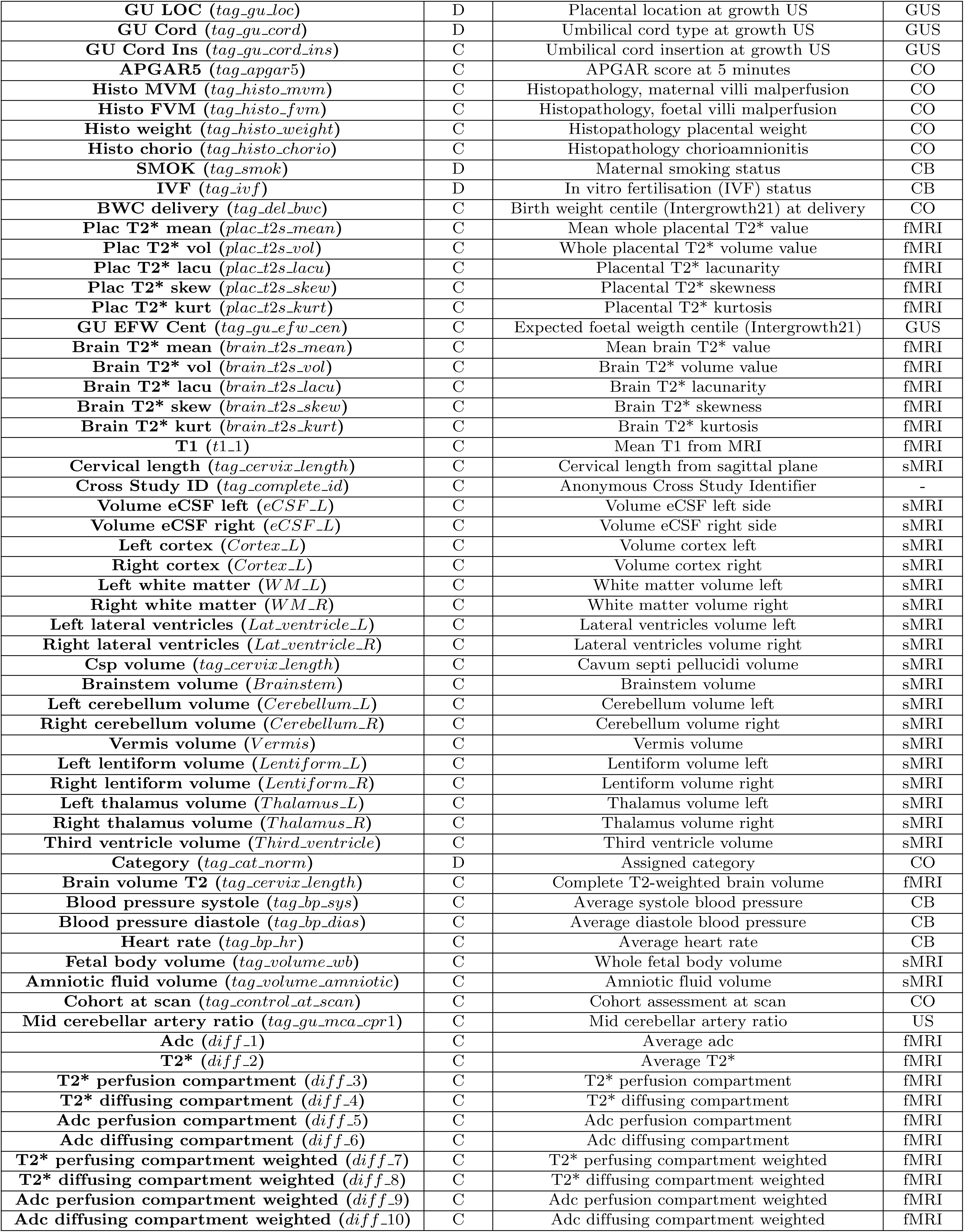
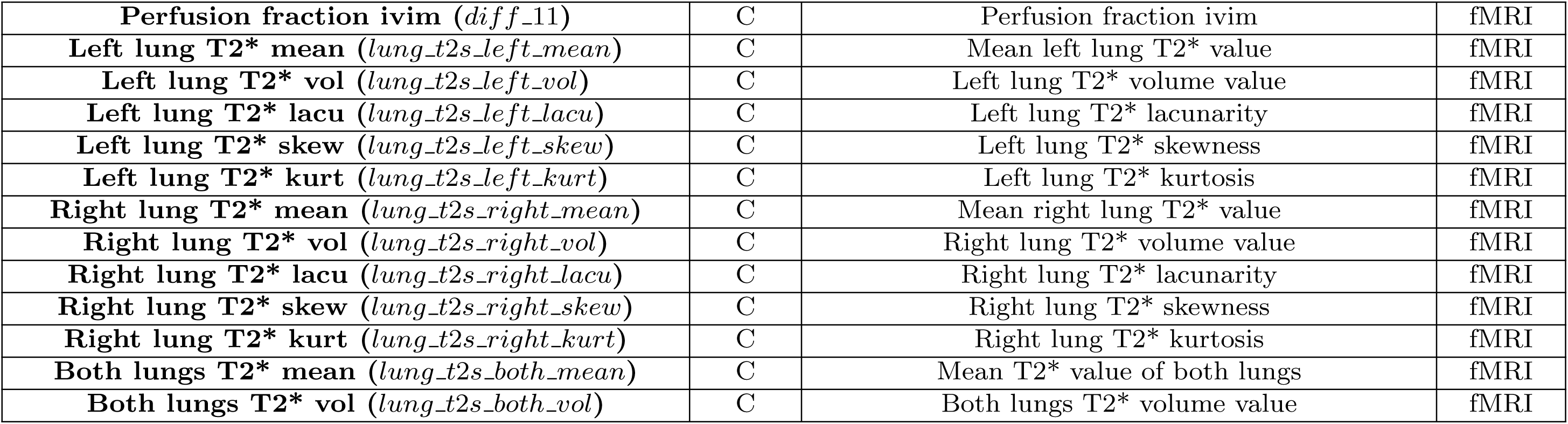
Description of the features. Features and outcomes available in the original data set. The first column is the name of the feature, the second column its type (C = continuous, Cat = categorical, D = discrete), the third column provides a short description, and the last column registers how each feature was acquired: clinical background (CB), clinical outcome (CO), structural MRI (sMRI), functional MRI (fMRI), growth ultrasound (GUS), and anomaly ultrasound (AUS).

**S2 Table.** Statistical summary of the features. Statistics of the data set after the first preprocessing steps and before imputation.

| Feature name | Missing | Mean | Median | STD | Skewness |
| --- | --- | --- | --- | --- | --- |
| *tag_ga* | 0 (0.0%) | 28.18 | 28.14 | 4.56 | -0.11 |
| *tag_typ* | 23 (9.47%) | 158.21 | 99.0 | 300.93 | 2.6 |
| *tag_scanner* | 0 (0.0%) | 1.0 | 1.0 | 0.0 | 0.0 |
| *tag_age* | 10 (4.12%) | 34.18 | 34.3 | 4.7 | -0.27 |
| *tag_bmi* | 21 (8.64%) | 23.77 | 23.65 | 2.98 | 0.48 |
| *tag_loc* | 35 (14.4%) | 3.53 | 2.0 | 2.09 | 1.76 |
| *tag_gadel* | 0 (0.0%) | 37.37 | 38.86 | 4.37 | -1.73 |
| *tag_mod* | 0 (0.0%) | 4.34 | 4.0 | 2.48 | 0.14 |
| *tag_sex* | 8 (3.29%) | 1.57 | 2.0 | 0.52 | 0.19 |
| *tag_bwg* | 7 (2.88%) | 2832.48 | 3062.5 | 917.24 | -0.91 |
| *tag_bwc* | 107 (44.03%) | 45.75 | 43.06 | 29.52 | 0.17 |
| *tag_parity* | 0 (0.0%) | 0.48 | 0.0 | 0.78 | 2.14 |
| *tag_prev_ptb* | 0 (0.0%) | 0.11 | 0.0 | 0.33 | 2.84 |
| *tag_bpd* | 122 (50.21%) | 73.51 | 74.0 | 12.86 | -0.29 |
| *tag_bpd_cent* | 124 (51.03%) | 49.3 | 47.0 | 28.02 | 0.05 |
| *tag_ce_tcd* | 123 (50.62%) | 33.88 | 32.55 | 8.42 | 0.08 |
| *tag_ce_tcd_cent* | 129 (53.09%) | 51.94 | 52.5 | 23.59 | -0.08 |
| *tag_post_hor_diam* | 125 (51.44%) | 6.42 | 6.35 | 1.61 | 0.22 |
| *eCSF_L* | 100 (41.15%) | 31941.45 | 33049.6 | 11969.29 | 0.17 |
| *eCSF_R* | 100 (41.15%) | 30944.74 | 31214.5 | 10941.05 | 0.35 |
| *Cortex_L* | 100 (41.15%) | 21351.56 | 16914.0 | 12237.17 | 0.74 |
| *Cortex_R* | 100 (41.15%) | 21422.58 | 17092.2 | 11960.04 | 0.73 |
| *WM_L* | 100 (41.15%) | 44681.85 | 41335.1 | 20056.3 | 0.35 |
| *WM_R* | 100 (41.15%) | 44771.93 | 41127.5 | 20324.06 | 0.37 |
| *Lat_ventricle_L* | 100 (41.15%) | 2154.67 | 2018.12 | 963.81 | 1.01 |
| *Lat_ventricle_R* | 100 (41.15%) | 2008.0 | 1891.96 | 893.35 | 0.89 |
| *CSP* | 100 (41.15%) | 497.36 | 471.24 | 224.35 | 0.85 |
| *Brainstem* | 100 (41.15%) | 3514.29 | 3428.88 | 1474.82 | -0.16 |
| *Cerebellum_L* | 100 (41.15%) | 2999.35 | 2493.24 | 1914.44 | 0.5 |
| *Cerebellum_R* | 100 (41.15%) | 2977.61 | 2348.0 | 1992.55 | 0.47 |
| *Vermis* | 100 (41.15%) | 936.0 | 831.04 | 535.0 | 0.25 |
| *Lentiform_L* | 100 (41.15%) | 2120.58 | 1953.5 | 1059.44 | 0.23 |
| *Lentiform_R* | 100 (41.15%) | 2041.67 | 1871.62 | 1033.36 | 0.15 |
| *Thalamus_L* | 100 (41.15%) | 1642.67 | 1483.75 | 819.65 | 0.29 |
| *Thalamus_R* | 100 (41.15%) | 1590.87 | 1420.96 | 801.94 | 0.26 |
| *Third_ventricle* | 100 (41.15%) | 140.56 | 143.68 | 70.51 | -0.08 |
| *tag_diabetes* | 7 (2.88%) | 0.5 | 0.0 | 2.13 | 4.3 |
| *tag_cat_norm* | 75 (30.86%) | 1.45 | 1.0 | 0.52 | 0.45 |
| *tag_hc* | 64 (26.34%) | 33.3 | 34.0 | 3.03 | -2.23 |
| *tag_hcc* | 121 (49.79%) | 53.64 | 56.6 | 33.57 | -0.12 |
| *tag_garom* | 0 (0.0%) | 37.0 | 38.86 | 5.07 | -1.78 |
| *tag_gu_efw_cen* | 103 (42.39%) | 54.11 | 61.19 | 32.35 | -0.41 |
| *tag_vol_t2w_complete* | 100 (41.15%) | 143758.08 | 134099.98 | 60465.11 | 0.26 |
| *tag_cpnr* | 102 (41.98%) | 0.38 | 0.36 | 0.12 | 0.93 |
| *tag_histo_weight* | 131 (53.91%) | 417.62 | 453.0 | 130.33 | -0.55 |
| *tag_histo_mvm* | 132 (54.32%) | 0.22 | 0.0 | 0.41 | 1.4 |
| *tag_histo_fvm* | 132 (54.32%) | 0.02 | 0.0 | 0.13 | 7.35 |
| *tag_histo_chorio* | 133 (54.73%) | 0.38 | 0.0 | 0.49 | 0.49 |
| *tag_anom_pi_left* | 191 (78.6%) | 1.19 | 1.07 | 0.52 | 0.5 |
| *tag_anom_pi_right* | 191 (78.6%) | 1.22 | 1.12 | 0.56 | 1.19 |
| *tag_anom_ga* | 53 (21.81%) | 20.02 | 20.0 | 0.9 | -3.34 |
| *tag_anom_loc* | 35 (14.4%) | 3.53 | 2.0 | 2.09 | 1.76 |
| *tag_anom_cord* | 69 (28.4%) | 1.11 | 1.0 | 0.56 | 5.32 |
| *tag_anom_cord_ins* | 182 (74.9%) | 5.7 | 8.0 | 2.86 | -0.66 |
| *tag_anom_hc* | 58 (23.87%) | 172.3 | 171.8 | 9.81 | 1.51 |
| *tag_anom_ac* | 58 (23.87%) | 150.4 | 150.2 | 11.03 | 0.83 |
| *tag_anom_bpd* | 71 (29.22%) | 47.56 | 47.65 | 3.15 | -2.64 |
| *tag_anom_fl* | 58 (23.87%) | 31.72 | 31.7 | 2.53 | 0.23 |
| *tag_gu_ga* | 94 (38.68%) | 28.65 | 28.43 | 4.63 | -0.37 |
| *tag_gu_hc* | 98 (40.33%) | 259.63 | 265.3 | 44.46 | -0.89 |
| *tag_gu_ac* | 98 (40.33%) | 239.14 | 239.3 | 49.19 | -0.41 |
| *tag_gu_bpd* | 100 (41.15%) | 73.0 | 74.8 | 13.2 | -0.72 |
| *tag_gu_fl* | 97 (39.92%) | 52.53 | 53.25 | 10.87 | -0.68 |
| *tag_gu_pi* | 112 (46.09%) | 1.08 | 1.03 | 0.22 | 1.65 |
| *tag_gu_efw* | 103 (42.39%) | 1378.66 | 1229.5 | 727.21 | 0.53 |
| *tag_gu_mca_pi* | 133 (54.73%) | 1.85 | 1.84 | 0.36 | -0.12 |
| *tag_gu_mca_psc* | 138 (56.79%) | 43.83 | 42.7 | 13.47 | 0.71 |
| *tag_gu_mca_cpr* | 158 (65.02%) | 1.82 | 1.83 | 0.45 | 0.0 |
| *tag_gu_notch* | 153 (62.96%) | 1.1 | 1.0 | 0.5 | 5.23 |
| *tag_gu_pi_left* | 153 (62.96%) | 0.87 | 0.85 | 0.26 | 0.71 |
| *tag_gu_pi_right* | 153 (62.96%) | 0.87 | 0.86 | 0.26 | 0.91 |
| *tag_gu_edf* | 112 (46.09%) | 1.11 | 1.0 | 0.62 | 6.12 |
| *tag_gu_loc* | 110 (45.27%) | 3.61 | 2.0 | 2.22 | 1.53 |
| *tag_gu_cord* | 140 (57.61%) | 1.06 | 1.0 | 0.44 | 8.1 |
| *tag_gu_cord_ins* | 161 (66.26%) | 3.28 | 4.0 | 1.28 | -1.24 |
| *tag_apgar5* | 26 (10.7%) | 9.25 | 10.0 | 1.5 | -3.74 |
| *tag_smok* | 33 (13.58%) | 1.41 | 1.0 | 1.08 | 2.35 |
| *tag_ivf* | 25 (10.29%) | 1.87 | 2.0 | 0.46 | -2.15 |
| *tag_bp_sys* | 105 (43.21%) | 106.22 | 104.22 | 12.78 | 0.54 |
| *tag_bp_dias* | 105 (43.21%) | 65.36 | 64.06 | 10.72 | 0.53 |
| *tag_bp_hr* | 118 (48.56%) | 77.5 | 76.4 | 10.1 | 0.27 |
| *tag_volume_wb* | 220 (90.53%) | 763487.03 | 683058.59 | 313741.75 | 0.59 |
| *tag_volume_amniotic* | 220 (90.53%) | 460971.62 | 526734.02 | 242231.28 | -0.32 |
| *tag_control_at_scan* | 0 (0.0%) | 0.62 | 1.0 | 0.49 | -0.49 |
| *tag_gu_mca_cpr1* | 138 (56.79%) | 1.79 | 1.83 | 0.46 | -0.22 |
| *tag_del_bwc* | 7 (2.88%) | 47.22 | 48.04 | 31.14 | -0.0 |
| *plac.t2s_mean* | 0 (0.0%) | 57.92 | 59.69 | 19.11 | -0.01 |
| *plac.t2s_vol* | 0 (0.0%) | 366.89 | 340.31 | 173.32 | 0.77 |
| *plac.t2s_lacu* | 0 (0.0%) | 19.47 | 19.08 | 6.16 | 2.53 |
| *plac.t2s_skew* | 0 (0.0%) | 13.33 | 2.36 | 30.53 | 3.49 |
| *plac.t2s_kurt* | 0 (0.0%) | 1.34 | 0.68 | 1.89 | 2.37 |
| *lung.t2s_left_mean* | 159 (65.43%) | 65.26 | 62.16 | 18.45 | 1.21 |
| *lung.t2s_left_vol* | 159 (65.43%) | 13.88 | 11.08 | 8.71 | 1.4 |
| *lung.t2s_left_lacu* | 159 (65.43%) | 20.06 | 18.43 | 8.72 | 1.58 |
| *lung.t2s_left_skew* | 159 (65.43%) | 7.27 | 1.78 | 13.54 | 2.99 |
| *lung.t2s_left_kurt* | 159 (65.43%) | 0.99 | 0.58 | 1.16 | 1.81 |
| *lung.t2s_both_mean* | 155 (63.79%) | 67.06 | 62.62 | 19.26 | 1.17 |
| *lung.t2s_both_vol* | 155 (63.79%) | 31.95 | 26.15 | 20.13 | 1.2 |
| *tag_vol_body* | 100 (41.15%) | 1009.95 | 971.36 | 456.13 | 8.94 |
| *tag_cervix_length* | 26 (10.7%) | 30.7 | 31.48 | 10.39 | -0.71 |
| *brain.t2s_mean* | 101 (41.56%) | 165.0 | 174.29 | 42.69 | -0.56 |
| *brain.t2s_vol* | 101 (41.56%) | 128.08 | 109.09 | 72.89 | 1.32 |
| *brain.t2s_lacu* | 101 (41.56%) | 68.79 | 73.92 | 19.64 | -0.77 |
| *brain.t2s_skew* | 101 (41.56%) | 2.27 | 0.17 | 6.07 | 4.43 |
| *brain.t2s_kurt* | 101 (41.56%) | 0.91 | 0.73 | 0.67 | 1.56 |
| *lung_t2s_right_mean* | 158 (65.02%) | 67.97 | 63.24 | 19.56 | 1.07 |
| *lung_t2s_right_vol* | 158 (65.02%) | 19.64 | 15.85 | 13.1 | 1.15 |
| *lung_t2s_right_lacu* | 158 (65.02%) | 22.13 | 19.95 | 9.71 | 1.22 |
| *lung_t2s_right_skew* | 158 (65.02%) | 7.66 | 2.42 | 14.4 | 4.1 |
| *lung_t2s_right_kurt* | 158 (65.02%) | 1.14 | 0.82 | 1.19 | 2.08 |
| *t1_1* | 199 (81.89%) | 1003.34 | 1016.46 | 243.97 | -0.8 |
| *diff_1* | 140 (57.61%) | 55.75 | 57.47 | 19.34 | -0.22 |
| *diff_2* | 140 (57.61%) | 0.0 | 0.0 | 0.0 | 1.14 |
| *diff_3* | 140 (57.61%) | 0.0 | 0.0 | 0.0 | 2.41 |
| *diff_4* | 140 (57.61%) | 0.0 | 0.0 | 0.0 | 0.0 |
| *diff_5* | 140 (57.61%) | 0.01 | 0.0 | 0.01 | 3.3 |
| *diff_6* | 140 (57.61%) | 0.0 | 0.0 | 0.0 | 0.0 |
| *diff_7* | 140 (57.61%) | 52.51 | 52.21 | 32.07 | 0.11 |
| *diff_8* | 140 (57.61%) | 0.03 | 0.03 | 0.01 | 1.02 |
| *diff_9* | 140 (57.61%) | 76.01 | 75.25 | 22.84 | 0.99 |
| *diff_10* | 140 (57.61%) | 0.0 | 0.0 | 0.0 | 0.62 |
| *diff_11* | 140 (57.61%) | 0.33 | 0.3 | 0.17 | 0.68 |
| *tag_complete_id* | 0 (0.0%) | 2400682.76 | 1000187.0 | 2864221.51 | 1.98 |

**S3 Table.** Hyperparameters of the base models. Hyperparameters investigated by the grid search for each of the base models

| Random Forests |  | Support Vector Regression |  | XGBoost |  |
| --- | --- | --- | --- | --- | --- |
| Hyperparameter | Values | Hyperparameter | Values | Hyperparameter | Values |
| *max_depth* | [3, 5, 10, 20, 50, 100] | *C* | [0.001, 0.01, 0.1, 1, 10, 100, 1000] | *max_depth* | [3, 5, 7, 10, 20, 50] |
| *max_features* | ['auto', 'sqrt', 'log2'] | *gamma* | ['scale', 'auto'] | *learning_rate* | [0.01, 0.05, 0.1, 0.3, 0.5] |
| *n_estimators* | [5, 10, 20, 50, 100, 250] | *kernel* | ['rbf', 'poly', 'sigmoid', 'linear'] | *min_child_weight* | [1,3,5,7] |
|  |  | *epsilon* | [0.001, 0.01, 0.1, 0.5, 1] | *gamma* | [0.1, 0.5, 0.8, 2, 5, 10] |
|  |  | *degree* | [2, 3] | *colsample_bytree* | [0.3, 0.5, 0.7] |

**S1 Fig. Details of First Preprocessing Steps.** Number of subjects kept after each initial preprocessing step.

**S1 Algorithm.**
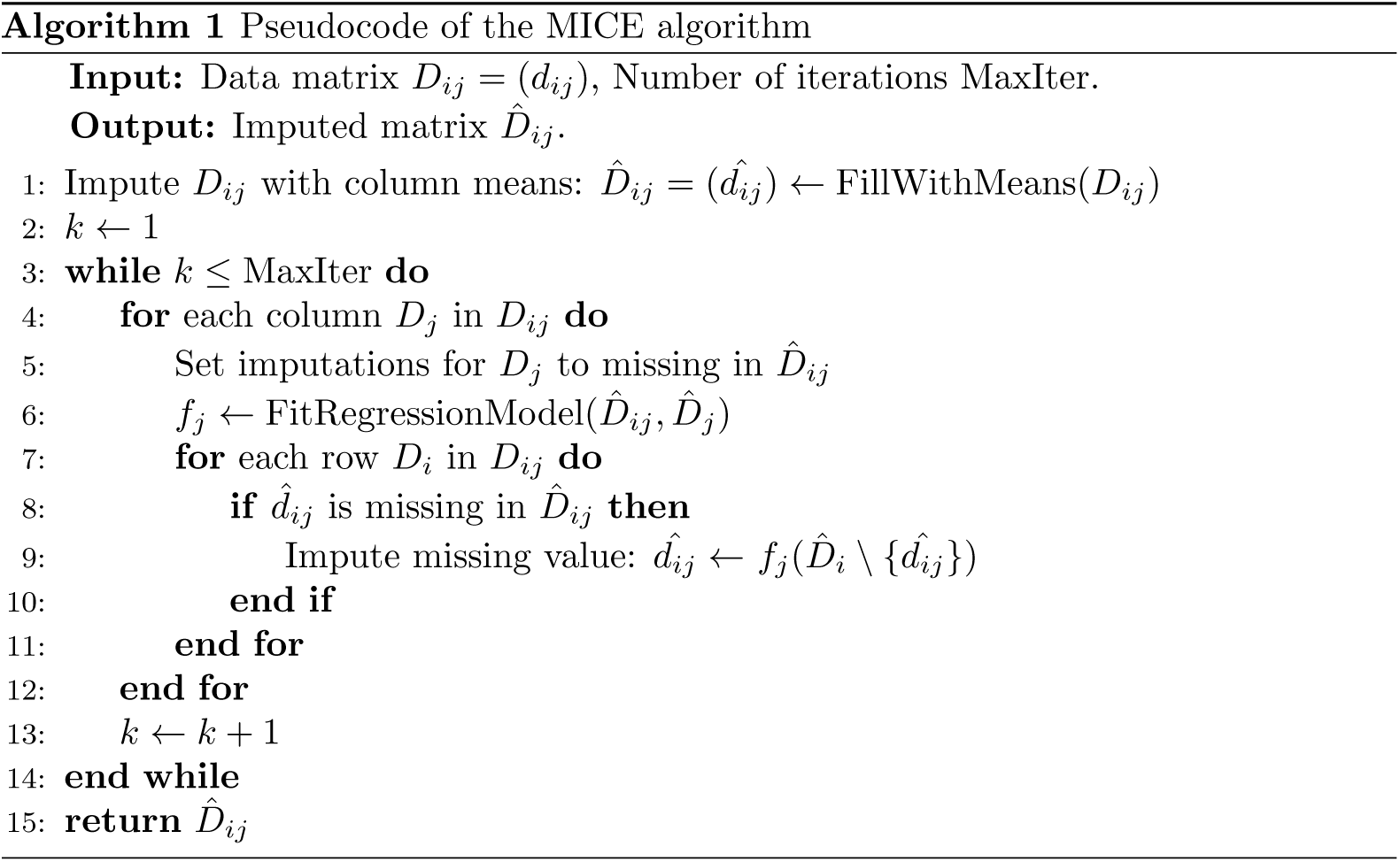
Pseudocode of the MICE algorithm.

## Notes

### Competing Interest Statement

The authors have declared no competing interest.

### Funding Statement

This work was supported by funding from the EPSRC Centre for Doctoral Training in Smart Medical Imaging (EP/S022104/1) to Diego Fajardo-Rojas, from
the Wellcome/EPSRC Centre for Medical Engineering[WT203148/Z/16/Z], a UKRI FLF [MR/T018119/1], and DFG Heisenberg funding through the High Tech Agenda Bavaria [502024488] to Jana Hutter, and from the NIHR Advanced Fellowship [NIHR3016640] and the MRC grant [MR/W019469/1] to Lisa Story. The funders had no role in software design, data collection or analysis, decision to publish, or preparation of the manuscript.

### Author Declarations

The data set used for this work comprises clinical records, MR data, and parameters manually extracted from ultrasound from 313 singleton pregnancies, acquired as part of 124 four ethically approved studies: 14/LO/1169 (Placenta Imaging Project, Fulham Research Ethics Committee, approval received September 23, 2016), 19-SS-0032 126 (Inflammation study in pregnancy, South East Scotland Ethics Committee, approval received March 7, 2019), 21/WA/0075 (Congenital Heart Imaging Programme, Wales Research Ethics Committee, approval received March 8, 2021), and 21/SS/0082 (Individualised Risk prediction of adverse neonatal outcome in pregnancies that deliver preterm using advanced MRI techniques and machine learning, South East Scotland Ethics Committee, approval received March 2022).

